# Cost of a new method of active screening for human African trypanosomiasis in the Democratic Republic of the Congo

**DOI:** 10.1101/2020.06.25.20139717

**Authors:** Rian Snijders, Alain Fukinsia, Yves Claeys, Alain Mpanya, Epco Hasker, Filip Meheus, Erick Miaka, Marleen Boelaert

**Author notes:** Corresponding author email Name: Rian Snijders, Postal address: Nationalestraat 155, 2000 Antwerp, Belgium.

## Abstract

**Background:** Human African trypanosomiases caused by the *Trypanosoma brucei gambiense* parasite is a lethal disease that killed thousands of people at the start of the 20^th^ century. Today, less than 1,000 cases are reported globally, and the disease is targeted for elimination and eradication. One of the main disease control strategies is active case-finding through outreach campaigns. In 2014, a new method for active screening was developed with mini, motorcycle-based, teams. This study aims to compare the cost of two approaches for active HAT screening, namely the traditional mobile teams and mini mobile teams.

**Methods:** We estimated annual economic costs for the two active HAT screening approaches from a health care provider perspective. Cost and operational data was collected for 12 months for 1 traditional team and 3 mini teams in the health districts of Yasa Bonga and Mosango in the Kwilu province of the Democratic Republic of the Congo. The cost per person screened and per person diagnosed was calculated. Univariate sensitivity analysis was conducted on important cost drivers.

**Results:** The study shows that the cost per person screened is lower for a mini team compared to a traditional team in the study setting (US$1.86 compared to US$2.08) as well as in a simulation analysis assuming both teams would operate in a setting with similar disease prevalence.

**Discussion:** Active HAT screening with mini mobile teams has a lower cost and could be a cost-effective alternative for active screening campaigns. Further research is needed to determine if mini mobile teams have similar or better yields than traditional mobile teams in terms of detections and cases successfully treated.

**AUTHOR SUMMARY:** Human African Trypanosomiasis (HAT) used to be a major public health problem in Sub-Saharan Africa, but the disease is becoming less frequent today as a result of sustained control efforts. Currently, the elimination of sleeping sickness is targeted as a public health problem by 2020 with interruption of transmission by 2030. To achieve these targets, a long-term commitment towards HAT control activities will be necessary with innovative disease control approaches accompanied by economic evaluations to assess their cost and cost-effectiveness in the changing context. Today, active case finding conducted through mass outreach campaigns accounts for approximately half of all identified cases in the Democratic Republic of the Congo. However, this strategy has become less efficient, with a dwindling “yield” in terms of the number of identified cases, translating to a higher cost per diagnosed HAT case. Therefore, different approaches to outreach campaigns need to be evaluated with a focus on reaching populations at risk for HAT.

This article presents the costs and outcomes of two approaches to active screening: traditional mobile teams and mini mobile teams.

This study shows that mini mobile teams could be a cost-effective alternative for active screening with a cost-per-person screened of US$1.86 compared to US$2.08. This approach could increase the screening coverage of populations at risk for HAT that are currently not being reached through the traditional approach. Future research is needed to evaluate the difference in HAT cases identified and treated by both approaches. This would allow a cost-effectiveness comparison of both strategies based on the cost-per-person diagnosed and treated.

## INTRODUCTION

Human African trypanosomiasis (HAT), or sleeping sickness, is a vector-borne disease believed to be invariably fatal when left untreated. There exist two forms of HAT, one caused by the Trypanosoma brucei (*T. b*.*) rhodesiense* and a second caused by the parasite *T. b. gambiense*. Infections with *T*.*b. gambiense* are responsible for more than 95% of the globally reported HAT cases and are the focus of this study. (1)

HAT is considered a public health problem because of the devastating epidemics in the 20^th^ century, but it is becoming more and more uncommon today thanks to sustained control efforts. (2) Therefore, the World Health Organization’s (WHO) Strategic and Technical Advisory Group for neglected tropical diseases decided to target the elimination of HAT as a public health problem by 2020 and interruption of transmission by 2030. (3) In 2018, 953 new HAT cases were declared globally, well below the targeted maximum of 2,000 cases. (4) The current method to control HAT is a combination of case-finding and treatment, and in some places, vector control as well. Case-finding is conducted either actively, through mass outreach campaigns by large mobile teams (here after called ‘traditional teams’) or passively in fixed health facilities. Currently, each of these strategies’ accounts for approximately half of all identified cases. Active case-finding has proven to be highly effective in poor, remote HAT endemic communities with limited access to health care facilities, but this strategy is labour-intensive, costly, and time-consuming as it generates a high opportunity cost for the populations screened because of the time they have to queue waiting for the service. (5, 6) In a context of near disease elimination, this control strategy also becomes less efficient, with a dwindling “yield” in the number of identified HAT cases, translating to a higher cost per detected case due to the decreasing prevalence and declining participation rates. Additionally, mass screening campaigns are characterised by heavy logistics-operations which limit the possibilities to organise a targeted and responsive screening in high-problem areas and in remote areas that are difficult to access by car. (7)

Five years ago, an alternative model for active HAT screening, called screening by “mini-teams”, was developed, which tries to mitigate the diminishing uptake and efficiency of traditional teams. (8) Qualitative research showed that communities prefer this type of screening because it is more adapted to their daily routine and guarantees more confidentiality, and therefore, they are also more likely to participate. (7, 9)

Only a few economic evaluations assess the cost and cost-effectiveness of HAT control activities, and they mainly focus on diagnostic algorithms for case detection, treatment options, and vector control. (6) If we want to achieve sustainable elimination of transmission, a longer-term commitment towards HAT control activities will be necessary, integrating improved tools and innovative disease control approaches. (10) This study aims to document the cost of two approaches to active HAT screening: traditional mobile teams and mini mobile teams, aiming to facilitate decisions on resource allocation for HAT control in different settings in the context of disease elimination.

## MATERIALS AND METHODS

### Study area

The study was conducted in two health zones in the former Bandundu province: Mosango and Yasa Bonga (Fig. 1). Both traditional and mini mobile teams have operated in these zones since 2016.

**Fig 1.**
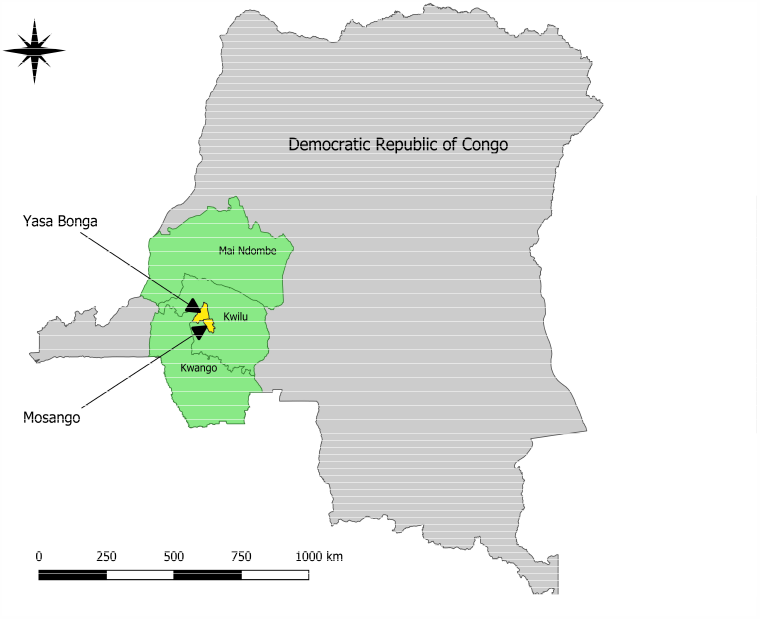
Location of the study area (map generated using QGIS 3.6.1) (11)

### Mobile screening strategies and diagnostic algorithms in DRC

A traditional mobile team consists of 8 to 9-members that travel from village to village using a truck. These teams invite the whole community to a centrally located open space in the village. A “mini-team” consists of only 4 people; three members of the team perform screening by visiting every family in a community on a door-to-door calling basis and the fourth team member performs parasitological confirmation of HAT suspects at a later moment. Both the traditional mobile team and mini team screen on average 300 people per day for 20 days a month for 11 months a year. The screening capacity of both types of teams is therefore estimated at 66,000 people annually.

#### Diagnostic algorithms

HAT diagnosis needs to be confirmed before a patient can be treated because of the toxicity and the complexity of the existing HAT treatment regimes. The disease is currently diagnosed through a combination of a serological test followed by more specific parasitological tests. The most frequently used serological test is the Card Agglutination Test for Trypanosomiasis (CATT), which is appropriate for mass population screening and distributed in vials of 50 tests. Once opened, the vials need to be used the same day, and specialised equipment (a rotator) requiring electricity and a cold chain for storage is needed. (12, 13) Recently, several rapid diagnostic tests (RDT) were developed and two are currently available on the market: RDT SD Bioline HAT from Standard Diagnostics and the HAT Sero-K-Set by Coris Bioconcept. These serological rapid tests have the advantage that they are produced for individual and mass use and do not require a cold chain or any other equipment. (13) During the study period, both the traditional team and mini teams performed the CATT test for screening. In July 2018 the mini teams started using RDTs for screening.

During the screening campaigns, the lymph nodes of all people with a positive CATT test and/or typical HAT symptoms (HAT suspects) were palpated. Upon detection of typically swollen lymph nodes, a lymph gland puncture (LGP) was performed and the fluid examined for parasites. HAT suspects without typical lymph nodes or with a negative lymph node examination were referred for microscopy tests in the following sequence: Capillary Tube Centrifugation (CTC) followed by the more sensitive Mini Anion Exchange Centrifugation Technique (mAECT). (14, 15) A HAT case was considered confirmed when one of the microscopy tests was positive (LGP, CTC, or mAECT). While traditional teams followed PNLTHA guidelines, mini mobile teams did not perform the CTC since their main energy source is 12-volt batteries charged through solar panels. No suitable 12-volt haematocrit centrifuges necessary for CTC could be found. Furthermore, 2 centrifuges would be too cumbersome on a motorcycle. (14)

#### Disease staging, monitoring, and treatment

HAT evolves in two stages: a haemo-lymphatic stage followed by a meningo-encephalitic stage when the parasite penetrates the blood-brain barrier and affects the central nervous system. During the study, the WHO guidelines stated a different treatment for each stage, requiring staging of disease through a lumbar puncture (LP) once the presence of the parasite was confirmed. (1, 16) First-stage HAT cases were treated with pentamidine and second-stage patients with nifurtimox-eflornithine combination therapy (NECT). (5) In August 2018, WHO published new guidelines for the treatment of sleeping sickness following the approval of the oral medicine fexinidazole, which is used for both stages. (17)

Traditional teams performed the LP on the spot and usually carry pentamidine for stage-one treatment at the nearest health centre. Stage-two patients were referred to the nearest hospital because NECT treatment requires intravenous infusions and close clinical monitoring.

Contrary to traditional teams, mini teams are not equipped for staging. Therefore, mini teams refer all confirmed HAT cases to hospitals for staging and treatment. In addition, traditional teams performed serial dilutions of CATT on HAT suspects with negative microscopy tests. People testing positive on CATT 1/8 are considered ‘serological cases’, to be staged and treated for HAT, like cases detected through LGP, CTC, or mAECT. People testing negative on CATT 1/8 were referred for monitoring by the local health centre.

Table 1 provides an overview of the two mobile screening strategies that were examined in this study.

**Table 1.** Screening strategies compared: a traditional mobile team and a mini mobile team

|  | **Traditional Mobile Team** | **Mini Mobile Team** |
| --- | --- | --- |
| Human Resources | 8 people | 4 people |
| Vehicle | One 4x4 vehicle | 4 motorcycles |
| Screening period | 12 months | 12 months |
| Serological test | CATT | CATT |
| Parasitological tests | LGP, CTC, mAECT, | LGP, mAECT |
| Monitoring test | CATT 1/8 | Ø |
| Stage determination | LP on site | LP in Health structure equipped to perform HAT microscopy & 2<sup>nd</sup> stage treatment |
| Treatment stage 1 | Nearby health centre | Nearby health centre |
| Treatment stage 2 | Health structure equipped to perform HAT microscopy & 2<sup>nd</sup> stage treatment | Health structure equipped to perform HAT microscopy & 2<sup>nd</sup> stage treatment |

CATT = Card agglutination test for trypanosomiasis; LGP = Lymph Gland Puncture; CTC = capillary tube centrifugation; mAECT = mini Anion Exchange Centrifugation Technique; LP = Lumbar Puncture

### Costing methodology

The study adopted the perspective of the PLNTHA and the public health care system. Data on resource consumption and prices were collected prospectively between May 2017 and April 2018 and complemented with financial records from the HAT control programme. Costs incurred by households were excluded; research costs for activities that were relevant to the implementation of the interventions were expressed in equivalent local costs. Costs were categorised as recurrent or capital (defined as equipment with a useful life of more than one year). Both financial and economic costs were estimated. Financial costs represent the actual quantities consumed and prices paid for consumables, including transportation costs during the study period as well as any durable equipment that was purchased specifically for these activities. Economic costs were estimated as the value of resources foregone that could have been used in other activities (“opportunity costs”). For capital equipment, the purchase or replacement value was considered and annualised based on the expected useful life and discounted at a discount rate of 3%.

For each approach, the annual cost was divided by the number of people screened for 12 months to calculate the annual cost per person screened. All costs exclude value-added taxes (VAT) since the PNLTHA and its main donors are VAT-exempt in the DRC (DRC VAT rate is 16%). (18) For items shipped to the DRC, the price was increased by 10% to account for the average shipment cost of goods between Europe and the DRC. All costs were recorded in the currency they were incurred in and converted to US$ following the average exchange rate of the study period (EUR to dollar: 1,18; CDF to dollar: 0,00065).

### Scenario analysis: adjusting for differences in contexts

We examined the degree to which the different contexts in which the traditional and mobile teams operate would have an impact on the observed cost differences. Because the data are incompatible with a traditional econometric analysis to control for differences in the background epidemiological context, we performed a small simulation study of the costs incurred by each team if they were to operate in populations that are epidemiologically identical.

In our modelled scenario, we assumed that both types of teams operated in the same context have a similar percentage of serological suspects based on the results in the study area but that each team uses their regular diagnostic algorithm (Traditional team: LGP, CTC, mAECT, CATT 1/8; Mini team: LGP, CTC, mAECT). Additionally, one-way sensitivity analysis was performed to consider the specific contribution, after control for background epidemiology, of the specificity and use of serological screening tests, the prevalence of the disease, performance of the mobile teams, impact of changes in the useful life of vehicles and motorcycles (maximum and minimum according to WHO Choice guidelines in Africa), discount rate, other important cost drivers such as the fuel cost and the price of mAECT, and the use of RDT’s as serological test. The diagnostic test and epidemiological parameters used for this scenario and the sensitivity analysis can be found in the supplementary information (SI_table1).

## RESULTS

### Operational results

Between May 2017 and April 2018, the traditional team screened 65,190 people, performed microscopy tests on all 276 HAT suspects identified, and diagnosed 11 new HAT cases. The mini teams screened on average 66,480 people per team, performed microscopy tests on 95% of 833 HAT suspects, and diagnosed 6 new HAT cases (Table 2). The percentage of positive CATT tests was higher for the mini teams compared to the traditional teams (1.4% vs. 0.4%) leading to more parasitological confirmation tests that had to be performed by the mini teams. None of the CATT 1/8 tests performed by the traditional team on serological suspects were positive.

**Table 2.** Number of tests performed, and HAT cases identified between May 2017 and April 2018

| Type of team | CATT | CATT + | LNA | CTC | mAECT | HAT stage 1 | HAT stage 2 |
| --- | --- | --- | --- | --- | --- | --- | --- |
| Traditional Mobile Team | 65,190 | 276 (0,42%) | 17 | 220 | 271 | 4 | 7 |
| Average Mini Mobile Teams | 66,480 | 833 (1.25%) | 137 | - | 795 | 2 | 4 |
| Mini Mobile Team 1 | 67,715 | 839 | 242 | - | 811 | 1 | 1 |
| Mini Mobile Team 2 | 61,958 | 806 | 60 | - | 772 | 5 | 10 |
| Mini Mobile Team 3 | 69,767 | 855 | 108 | - | 801 |  |  |
| **Average all Mobile Teams** | **65,835** | **555** | **77** | **110** | **533** | **3** | **5** |
| **Total all Mobile teams** | **264,630** | **2,776 (1.05%)** | **427** | **220** | **2,655** | **10** | **18** |

### Financial and economic costs

#### Financial costs

The average annual financial cost for a traditional team was estimated at 177,000 $ (between 163,731 and 228,191) and for a mini team around 140,000 $ (between 130,617 and 171,926). The annual variations can be explained by the expenses related to the replacement of capital equipment (SI_Table 14).

#### Economic costs

Table 3 shows the economic costs per item for both approaches, the overall annual and the cost per person screened/diagnosed. The cost per person screened by a mini team (1.86$) was 12% lower than for a traditional team (2.08$) while the cost per person diagnosed is 44% lower for a traditional team (12,302$) compared to a mini team (21,893$). Detailed information per cost item is available in SI_Tables 2-13.

**Table 3.** Annual economic costs of active screening in Yasa Bonga and Mosango, by screening approach

|  | Traditional Team |  | Mini team |  |
| --- | --- | --- | --- | --- |
| | Cost (US\$) | % | Cost (US\$) | % |
| **Capital costs** | **10,782** | **8.0** | **8,582** | **6.9** |
| Vehicle/motorcycles | 5,073 | 3.7 | 4,136 | 3.3 |
| Medical and laboratory equipment | 1,944 | 1.4 | 1,147 | 0.9 |
| Energy source | 344 | 0.3 | 419 | 0.3 |
| Electronics | 624 | 0.5 | 554 | 0.4 |
| Other equipment | 973 | 0.7 | 504 | 0.4 |
| Training | 1,823 | 1.3 | 1,823 | 1.5 |
| **Recurrent costs** | **124,540** | **92.0** | **114,999** | **93.1** |
| Human Resources | 31,074 | 23.0 | 20,892 | 16.9 |
| Laboratory & medical supplies |  |  |  |  |
| Screening tests (CATT & Discarded CATT) | 53,587 | 39.6 | 58,355 | 47.2 |
| Parasitological confirmation | 1,793 | 1.3 | 4,329 | 3.5 |
| Staging | 208 | 0.2 | 0 | 0.0 |
| Surveillance | 932 | 0.7 | 0 | 0.0 |
| Other supplies and materials | 5,615 | 4.1 | 5,045 | 4.1 |
| Fuel costs | 5,719 | 4.2 | 2,299 | 1.9 |
| Management | 25,611 | 18.9 | 24,080 | 19.5 |
| **Total screening cost** | **135,322** | **100** | **123,582** | **100** |
| Cost per person screened | 2.08 |  | 1.86 |  |
| Cost per person diagnosed with HAT | 12,302 |  | 21,893 |  |

### Scenario analysis: costs in identical settings

Table 4 shows the cost per person screened for a theoretical scenario in which both types of teams operate in an identical context: 2.14$ for a traditional team and 1.86$ for a mini team. This cost is slightly higher for the traditional team than observed in Yasa Bonga and Mosango. In the model, the number of CATT positives is estimated based on the average number of CATT positives detected during the study (1.05%). The mini teams observed around 3 times more CATT positive tests than the traditional teams. This results in a higher number of CATT positives in the model for the traditional team and therefore a higher cost for parasitological confirmation and surveillance tests than observed in the study setting. The overall cost per person screened by a mini team is 0.29$ or 15% lower than by a traditional team. Over 80% of this difference can be explained by the lower costs for human resources and means of transportation (motorcycles and fuel consumption) of a mini team.

**Table 4.** Results scenario analysis: costs of a traditional and mini team in an identical setting

|  | Traditional Team |  | Mini team |  | Difference |  |
| --- | --- | --- | --- | --- | --- | --- |
| | \$ | % | \$ | % | \$ | % |
| **Capital equipment** | **10,782** | **7.6** | **8,582** | **7.0** | **0.03** | **11.6** |
| **Annual recurrent costs** | **130,751** | **92.4** | **114,039** | **93.0** | **0.25** | **88.4** |
| Human resources | 31,074 | 22.0 | 20,892 | 17.0 | 0.15 | 53.8 |
| Laboratory & medical supplies | 61,922 | 44.8 | 61,849 | 50.4 | 0.00 | 0.4 |
| Screening tests (CATT & discarded CATT) | 54,253 | 38.3 | 57,934 | 47.2 | -0.06 | -19.5 |
| Parasitological confirmation | 5,140 | 3.6 | 3,915 | 3.2 | 0.02 | 6.5 |
| Staging | 114 | 0.1 | - | 0.0 | 0.00 | 0.6 |
| Surveillance | 2,415 | 1.7 | - | 0.0 | 0.04 | 12.8 |
| Other supplies and materials | 5,615 | 4.0 | 5,045 | 4.1 | 0.01 | 3.0 |
| Fuel costs | 5,719 | 4.0 | 2,299 | 1.9 | 0.05 | 18.1 |
| Management | 26,421 | 18.7 | 23,955 | 19.5 | 0.04 | 13.0 |
| **Total screening** | **141,533** | **100** | **122,621** | **100** |  |  |
| Cost per person screened | 2.14 |  | 1.86 |  | 0.29 | 15.4 |
| Cost per person diagnosed with HAT | 23,551 |  | 22,888 |  |  |  |

### One-way sensitivity analysis of cost drivers

Figure 2 shows the impact on the cost per person screened when changing the variables used for the modelled scenario individually (SI_Table 16-24). For both approaches, the main cost drivers were the unit cost and the specificity of the serological test. Assuming that the CATT has a high specificity (0.993) has a small impact on the cost per person screened because of the high number of cases detected among the CATT positive tests in the study area (28/2,776). (21) In a context of disease elimination with very low prevalence, a lower specificity of the serological test will result in a higher number of false positives and therefore more microscopy tests. Using serological tests with a higher or lower specificity has a big impact on the cost per person screened.

**Fig 2.**
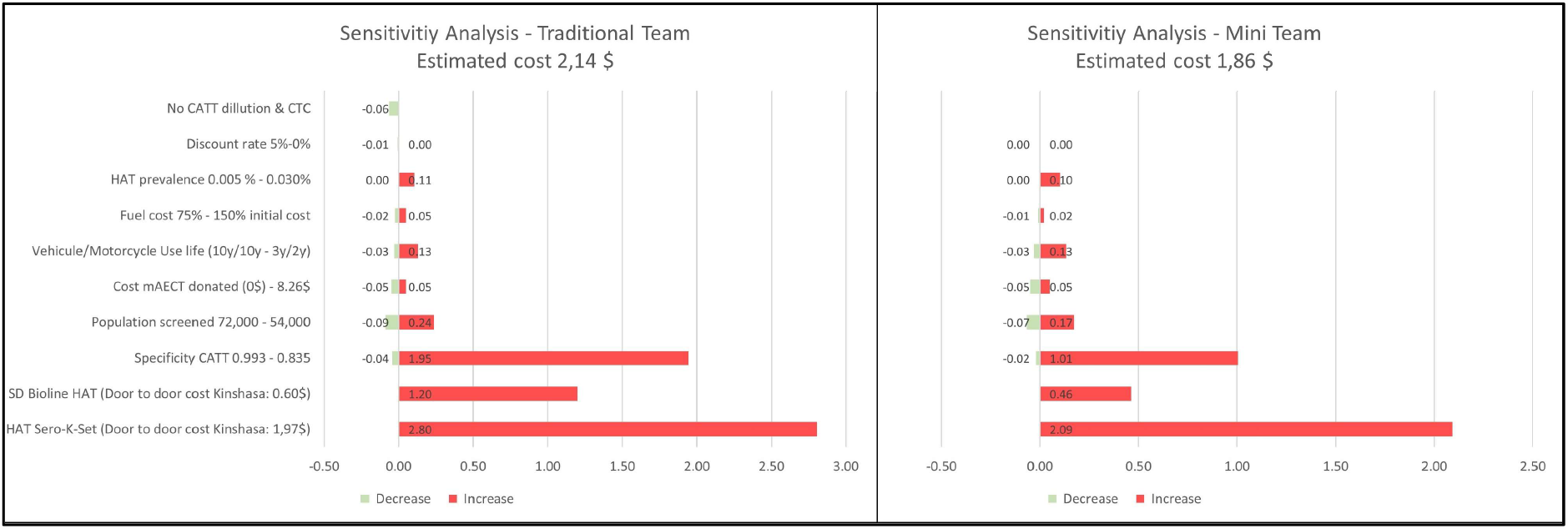
One-way sensitivity analysis – Additional cost/saving per person screened

Since the beginning of 2018, the mini mobile teams use HAT RDTs for active screening. The SD Bioline HAT RDT (0.60$) is less expensive than the CATT (0.74 $) but includes a subsidy of 0.25$ per RDT produced paid externally to the supplier. (19) The HAT Sero-K-Set RDT (1.97$) is much more expensive. The literature reports a lower specificity for both RDT’s than for the CATT. Therefore, the use of both RDT’s will push the cost per person screened upwards. When using the more expensive RDT (HAT Sero-K-Set) the cost per person screened could almost double for both approaches (4.95$ VS 2.14$; 3.95$ VS1.86$). (20)

During the study period, the diagnostic algorithm of a traditional team included 2 tests (CTC and CATT 1/8) that were not performed by the mini team. Excluding these tests from the traditional team’s diagnostic algorithm would lower their cost per person by 0.06$ to 2.08$ per person screened.

The study considered the purchase price of the mAECT, but currently, the gel needed to produce this test is donated. This sensitivity analysis also looked at the impact if this gel would no longer be donated. The cost per person screened would increase and the impact depends on the specificity of the serological tests.

The remaining variables (HAT prevalence, fuel cost, useful life of vehicles and motorcycles) have a much smaller impact on the cost per person screened.

## DISCUSSION

The current paper is a contribution to cost HAT efforts in the DRC. Past estimates of costs are outdated and innovative methods currently available were not considered. This study demonstrates that the new active screening approach by mini teams costs less than screening with traditional teams. The results are valid in the study area (1.86 $ vs 2.08 $) as well as for a scenario analysis assuming both approaches are implemented in an identical setting (1.86 $ vs 2.14$). A costing study in 2007 reported a higher cost per person screened by a traditional team in context with a higher prevalence, namely between 1.96 € and 2,99 € (or 2.7 $ and 4.1 $ in 2018). (14) A cost-effectiveness analysis comparing the societal costs (including the costs for the patient) and outcomes for active screening by traditional mobile teams using CATT or RDT estimated the cost per person screened with CATT at 2.31 $ (VS 2.14$) and with RDT at 2.37 $ (VS 3.34$). The differences can mainly be explained by the inclusion of the patient costs and the differences in parasitological confirmation tests due to the use of a much lower specificity of the CATT and a higher specificity for the SD Bioline HAT. (19) Research showed that people at risk for HAT are more likely to participate in screening activities by a mini team as they are contacted in person, do not need to queue, the moment of screening can be adapted to their daily routines and their privacy is respected. (9) Additionally, mini teams could reach areas inaccessible by vehicle, and investment and fuel costs of mini teams are much lower than for a traditional team, making them more suitable to be deployed in remote areas, regions inaccessible by car or to boost active screening activities for a short period.

A disadvantage in the current set up of mini teams is that they have difficulties ensuring that all HAT suspects undergo parasitological because such confirmation usually takes place at least 1 or 2 weeks later. Traditional teams, on the other hand, perform screening and confirmation simultaneously, ensuring continuity of care. The delay between screening and confirmation for mini teams could be resolved by making the screeners and the microscopist move around together but then additional equipped microscopists might be needed which would increase the cost per person screened. The problem would also be resolved if Fexinidazole or Acoziborole, a one dose drug against both stages and currently in the clinical trial phase, would be safe enough to treat serological HAT suspects making routine parasitological confirmation obsolete. (22) Additionally, mini teams and traditional teams use a different diagnostic algorithm (use of CTC and CATT dilution). It would make sense considering stopping using both additional tests in a context of low prevalence if they do not increase the sensitivity of the diagnostic algorithm because they increase the cost per person screened significantly. (23)

Currently, the mini mobile teams are using HAT RDTs, therefore their cost for active screening is most likely between 25% or even 115% higher than reported in this study, depending on the RDT they are using due to the higher purchase cost of the serological tests and the lower specificity.

Overall HAT screening by mini teams could be a cost-efficient alternative for active screening if they have similar or better outcomes in terms of the detection rate and enrolment in treatment. Better accessibility to populations at risk, the sensitivity of the diagnostic algorithm, and the delay between serological and parasitological tests could affect the number of cases identified and the enrolment in treatment and therefore the effectiveness of the teams. This approach should be considered a valid alternative to the traditional way of active HAT screening, but further research is needed to evaluate the difference in HAT cases identified and treated. This would allow calculating the cost per person diagnosed and treated for both strategies and performing a cost-effectiveness comparison of both strategies.

## Data Availability

All relevant data are within the manuscript and its Supporting Information file.

## Supporting information

SI_SupplementaryInformation (pdf)

## Ethics approval and consent to participate

Ethical approval for this study was obtained from the Institutional Review Board (IRB) of the University of Antwerp, Belgium, as well as from the IRB at Ecole de santé Publique of the University of Kinshasa, RDC.

## Acknowledgements

The authors acknowledge the staff of the PNLTHA in the DRC for their contribution under difficult field conditions.

## Contributors

The study was conceptualised and designed by RS, YC, AM, EH, FM, MB. MB and EH acquired the funding for this study. Data collection was done by RS, AF. RS performed the data analysis and drafted the manuscript. AF, FM, and MB contributed to the data analysis. AF, YC, AM, EH, FM, MB critically revised several versions of the manuscript. All authors approved the final version of the manuscript before submission.

## Disclaimer

Where authors are identified as personnel of the International Agency for Research on Cancer/World Health Organization, the authors alone are responsible for the views expressed in this article and they do not necessarily represent the decisions, policy or views of the International Agency for Research on Cancer /World Health Organization

